# Nasal-Swab Testing Misses Patients with Low SARS-CoV-2 Viral Loads

**DOI:** 10.1101/2020.06.12.20128736

**Authors:** Cody Callahan, Rose A. Lee, Ghee Rye Lee, Kate Zulauf, James E. Kirby, Ramy Arnaout

**Author notes:** To whom correspondence should be addressed at Ramy Arnaout, MD, DPhil, Beth Israel Deaconess Medical Center, 330 Brookline Avenue—YA309, Boston, MA 02215.

## Abstract

The urgent need for large-scale diagnostic testing for SARS-CoV-2 has prompted pursuit of sample-collection methods of sufficient sensitivity to replace sampling of the nasopharynx (NP). Among these alternatives is collection of nasal-swab samples, which can be performed by the patient, avoiding the need for healthcare personnel and personal protective equipment.

Previous studies have reached opposing conclusions regarding whether nasal sampling is concordant or discordant with NP. To resolve this disagreement, we compared nasal and NP specimens collected by healthcare workers in a cohort consisting of individuals clinically suspected of COVID-19 and outpatients known to be SARS-CoV-2 RT-PCR positive undergoing follow-up. We investigated three different transport conditions, including traditional viral transport media (VTM) and dry swabs, for each of two different nasal-swab collection protocols on a total of 308 study participants, and compared categorical results and Ct values to those from standard NP swabs collected at the same time from the same patients. All testing was performed by RT-PCR on the Abbott SARS-CoV-2 RealTime EUA (limit of detection [LoD], 100 copies viral genomic RNA/mL transport medium). We found high concordance (Cohen’s kappa >0.8) only for patients with viral loads above 1,000 copies/mL. Those with viral loads below 1,000 copies/mL, the majority in our cohort, exhibited low concordance (Cohen’s kappa = 0.49); most of these would have been missed by nasal testing alone. Previous reports of high concordance may have resulted from use of assays with higher LoD (≥1,000 copies/mL). These findings counsel caution in use of nasal testing in healthcare settings and contact-tracing efforts, as opposed to screening of asymptomatic, low-prevalence, low-risk populations. Nasal testing is an adjunct, not a replacement, for NP.

## Introduction

Controlling the COVID-19 pandemic will require a massive expansion of testing for SARS-CoV-2 in several different clinical and epidemiological contexts. Until recently, nasopharyngeal (NP) swabs were the United States Centers for Disease Control and Prevention’s (CDC) preferred specimen type, as these specimens were thought to provide the most robust detection of patient infection. However, there are conflicting reports as to which of several specimen types bear the highest viral load^1–3^, and ultimately the “preferred-specimen” specification was removed from interim CDC guidance on 29 April 2020^4^. Sensitivity is a complex issue, however, as detection in the upper airways (nasopharynx and oropharynx) is affected by multiple factors including duration of illness prior to testing^5^ as well as the limit of detection (LoD) of the RT-PCR assay used^6^.

Availability of NP swabs and the resources to establish NP collection sites with specimen collection personnel have remained critical bottlenecks. To resolve these issues, healthcare systems have adopted multiple different strategies, including engaging industrial manufacturers to mass produce novel 3D-printed NP swabs^7^, as well as evaluating different specimen types and alternative sample-collection strategies^8–16^. Assessment of nasal swabs is a rapidly growing area of interest, specifically because this specimen type involves a less invasive procedure than NP swabs, as only the anterior-to-mid-turbinate area of the nasal passages is accessed. Accordingly, such samples can be self-collected by patients with a simple set of instructions, alleviating the need for highly trained medical personnel for specimen collection and reducing use of personal protective equipment (PPE) in short supply.

Many of the US Food and Drug Administration Emergency Use Authorization (FDA EUA) RT-PCR assays have approval for use of nasal swabs as a specimen type, but how well nasal swabs perform compared to NP swabs remains unclear. To date, nasal-swab studies have shown conflicting results, with some researchers reporting similar test performance to NP swabs and others finding decreased sensitivity^8,10,12–16^. Reconciling these differences is challenging, as these studies employed different sampling materials, collection methods, and RT-PCR assays. To address these conflicting reports, here we describe results of a six-arm, 308-subject study comparing two different healthcare-worker nasal-swab collection procedures and three different transport conditions, including in viral transport media (VTM) and dry transport. We discuss our findings in the context of prior reports (including preprints), to more systematically assess nasal-swab test performance and its potential role(s) in addressing diagnostic and epidemiologic needs in the COVID-19 pandemic.

## Materials and Methods

### Trial design

Participants were adults over 18 years of age tested for SARS-CoV-2 during the normal course of clinical care, based either on clinically suspected COVID-19 infection or follow-up after previous SARS-CoV-2-positive RT-PCR testing. Participants were asked to be swabbed twice, first with one of the nasal swabs under study (see below for swab-collection protocols) and then with a standard NP swab. To control for potential variability related to self-swabbing, sample collection was performed by trained nurses or respiratory-therapy staff (“study staff”) with training and oversight from the respiratory therapy department at Beth Israel Deaconess Medical Center (BIDMC) drive-through/walk-up (“drive-through”) COVID-19 testing sites. Individuals with known thrombocytopenia (<50,000 platelets/µl) were excluded from the study to avoid risk of bleeding. This study was reviewed and approved by BIDMC’s institutional review board (IRB protocol no. 2020P000451).

### Transport conditions and swabs used

Standard nasal swabs were compared under three different specimen-transport conditions: (*i*) a guanidine thiocyanate (GITC) transport buffer, part of the Abbott multi-Collect Specimen Collection Kit, catalog no. 09K12-004; Abbott Laboratories, Abbott Park, IL), (*ii*) dry, with no buffer; and (*iii*) in modified CDC viral transport media (VTM) (Hank’s balanced salt solution containing 2% heat inactivated FBS, 100µg/mL gentamicin, 0.5µg/mL fungizone, and 10mg/L Phenol red, produced by the Beth Israel Deaconess Medical Center [BIDMC] Clinical Microbiology Laboratories^17^). The nasal swab used was the included Abbot swab for the GITC arm and the Hologic Aptima Multitest Swab otherwise (catalog no. AW-14334-001-003; Hologic Inc. Marlborough, MA). The NP swab used was the Copan BD ESwab collection and transport system swab (catalog no. 220532; Copan Diagnostics Inc., Murietta, CA).

### PCR compatibility

Although all swabs are routinely used for PCR testing, as a double-check each swab type was assessed for PCR compatibility by overnight incubation in 3 mL of modified CDC VTM (allowing potential PCR inhibitors time to leech into media), spiking 1.5 mL of media with 200 copies/mL of control SARS-CoV-2 amplicon target (twice the LoD of our system), vortexed, and tested using the Abbott RealTime SARS-CoV-2 Assay on an Abbott m2000 RealTime System platform^18^, the assay and platform used for all testing in this report, following the same protocol used for clinical testing (see below). All swabs examined in this study passed this quality-control testing for lack of RT-PCR inhibition based on observation of Ct values within expected quality control limits^17^.

### Swab collection protocols (Fig. 1)

For Procedure 1, for each naris, the swab tip was inserted into the nostril, the patient was told to press a finger against the exterior of that naris, and the swab was rotated against this external pressure for 10 seconds (Fig. 1a); this procedure was repeated with the same swab on the other naris, and then the swab was placed into the collection tube for transport to the laboratory for testing. For Procedure 2, the swab was inserted into the naris until resistance was felt, and the swab was then rotated for 15 seconds without external pressure (Fig. 1b); this procedure was repeated with the same swab on the other naris, and the swab was then placed into the collection tube for transport^15^. The NP-swab sample was collected from a single naris by standard technique: insertion to appropriate depth, 10 rotations, removal, and placement into transport-media tube containing VTM^4^. To maximize collection of material from the nares, in all cases sampling using the nasal swab (both nares) was performed first, before the NP swab.

**Figure 1:**
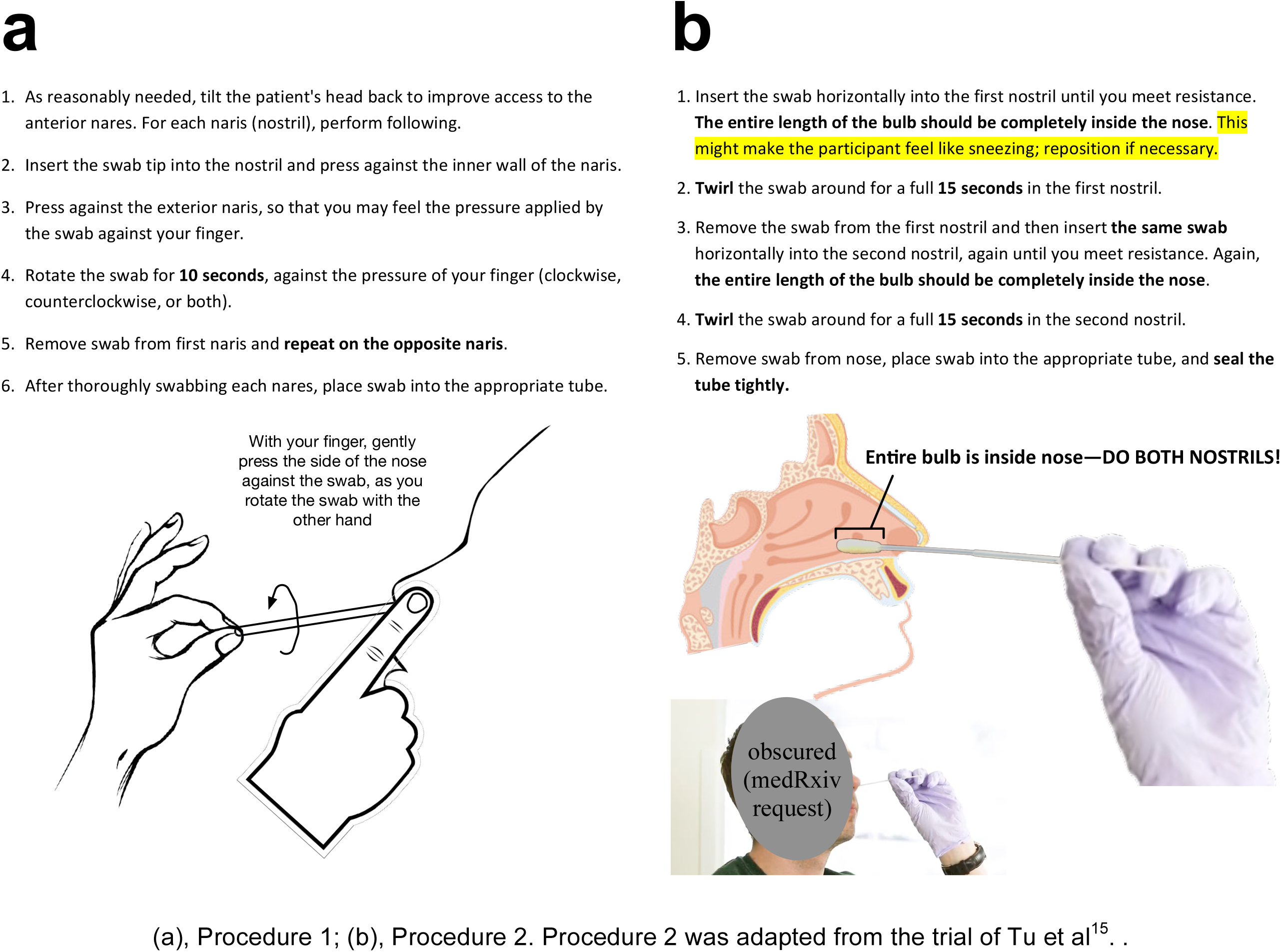
Nasal swabbing instructions

### Sample processing and testing

Samples were sent to the BIDMC Clinical Microbiology Laboratories for testing. Dry swabs were eluted in 2mL of Abbott *m*Wash1 (100mM Tris with guanidinium isothiocyanate (GITC) and detergent). Swabs transported in GITC buffer were supplemented with 1mL of Abbott *m*Wash1 solution to achieve minimum volume requirements for testing. Tests were performed with 1.5mL of sample media using the Abbott RealTime SARS-CoV-2 assay for EUA for use with nasopharyngeal and nasal swabs^18^. This dual-target assay detects both the SARS-CoV-2 RdRp (RNA-dependent RNA polymerase) and N (nucleocapsid) genes with an in-lab verified LoD of 100 copies/mL^6,17^.

### Statistical analyses

RT-PCR results reported categorically as either positive or negative, and these were used for concordance testing by Cohen’s kappa^19^.

For analyses based on cycle-threshold (Ct) values, for discordant samples (positive nasal-swab/negative NP-swab result or vice versa), the negative result was assigned a Ct value of 37, the total number of cycles run. Conversion to viral load was performed as described previously^6^.

We tested whether Ct values for a given set of nasal swabs differed from the Ct values for the paired NP swabs (the controls) using Wilcoxon’s paired *t*-test. This tested the null hypothesis that values for controls and prototypes are drawn from the same underlying distribution; p>0.05 was interpreted as no difference. We used bootstrapping to test whether the *n* results for a given arm exhibited appreciable differences from others, specifically by testing whether a given arm differed from random samples from (*i*) results pooled across the three arms that used the same nasal-swab sampling procedure or (*ii*) all results. For each bootstrap test, we sampled *n* data points at random from the larger pool to create a synthetic dataset, calculating Cohen’s kappa on this synthetic dataset, and repeating this process for 100,000 synthetic datasets to create a distribution (histogram) of kappa values; this distribution constitutes a null model of the kappa one would expect to observe by chance in a sample of *n* results, given the data in the larger pool. Using this distribution, we then calculated the probability of observing a kappa at least as high as the kappa actually observed for the *n* results from a given arm, to test for consistency with expectation; inconsistency (p<0.05 or *p*>0.95) would reject the null hypothesis that the study arm and the larger pool are statistically indistinguishable (as measured by kappa). For completeness, we performed the same bootstrap analysis to compare Procedure 1 results to all results and to compare Procedure 2 results to all results.

We used Python (v3.6-3.8) and its NumPy, SciPy, Matplotlib, Pandas, and ct2vl libraries for the above analyses and related visualizations.

### Literature review

We searched Pubmed and the preprint servers bioRxiv and medRxiv through June 1, 2020 for all literature on nasal-swab sampling for SARS-CoV-2 and extracted sample sizes, collection methods, RT-PCR assay information, and 2×2 contingency table data comparing nasal swabs to NP swabs wherever available.

## Results

Table 1 shows the numbers of patients tested in each of the six arms of our nasal-vs. NP-swab study. Visual inspection of plots of the Ct values of the nasal swab vs. NP-swab controls suggested worse performance for nasal swabs across all six arms, with no obvious differences between the two swab procedures or among the dry-swab, VTM, or GITC collection methods (Fig. 2). Statistical testing confirmed that results for each arm were indistinguishable from the overall results, supporting the functional equivalence of all swab/transport-condition combinations (Table 2). For concordant positives, comparison of Ct values between nasal and NP swabs showed higher Cts for nasal swabs than for NP swabs, suggesting slightly but consistently lower yield from the nasal swabs (Wilcoxon *p*=9×10^−11^). Consistent with this conclusion, there was a marked increase in false negatives for NP-swabs with higher Ct values (lower viral loads), resulting in low concordance overall (Cohen’s kappa=0.49) (Fig. 2).

**Table 1:** Number of specimens by study arm. Procedures 1 and 2 as in Methods and Fig. 1. Numbers in parentheses indicate study arms.

| Collection<br>method | Transportation conditions |  |  |
| --- | --- | --- | --- |
|  | GITC | Dry | VTM |
| Procedure 1 | 47 (1) | 36 (2) | 39 (3) |
| Procedure 2 | 65 (4) | 61 (5) | 60 (6) |

**Table 2:** Comparability of study arms and subsets. Listed are *p*-value for bootstrap comparison of linear-regression fits of Ct values of nasal swab vs. NP swab for observed samples for arms 1-6 from Table 1 vs. 10,000 random samples of either Procedure 1 (top three rows), Procedure 2 (next three rows), total (next six rows). Bottom two rows are bootstrap comparisons of all Procedure 1 (i.e., arms 1-3) to total and Procedure 2 (arms 4-6) to total. All comparisons show *p*>0.05, interpreted as no significant differences among arms and procedures.

| Comparison | p-value |
| --- | --- |
| Arm (1) vs. all Procedure 1 | 0.11 |
| Arm (2) vs. all Procedure 1 | 0.85 |
| Arm (3) vs. all Procedure 1 | 0.62 |
| Arm (4) vs. all Procedure 2 | 0.78 |
| Arm (5) vs. all Procedure 2 | 0.30 |
| Arm (6) vs. all Procedure 2 | 0.53 |
| Arm (1) vs. total | 0.27 |
| Arm (2) vs. total | 0.82 |
| Arm (3) vs. total | 0.62 |
| Arm (4) vs. total | 0.71 |
| Arm (5) vs. total | 0.23 |
| Arm (6) vs. total | 0.44 |
| Procedure 1 vs. total | 0.66 |
| Procedure 2 vs. total | 0.29 |

**Figure 2:**
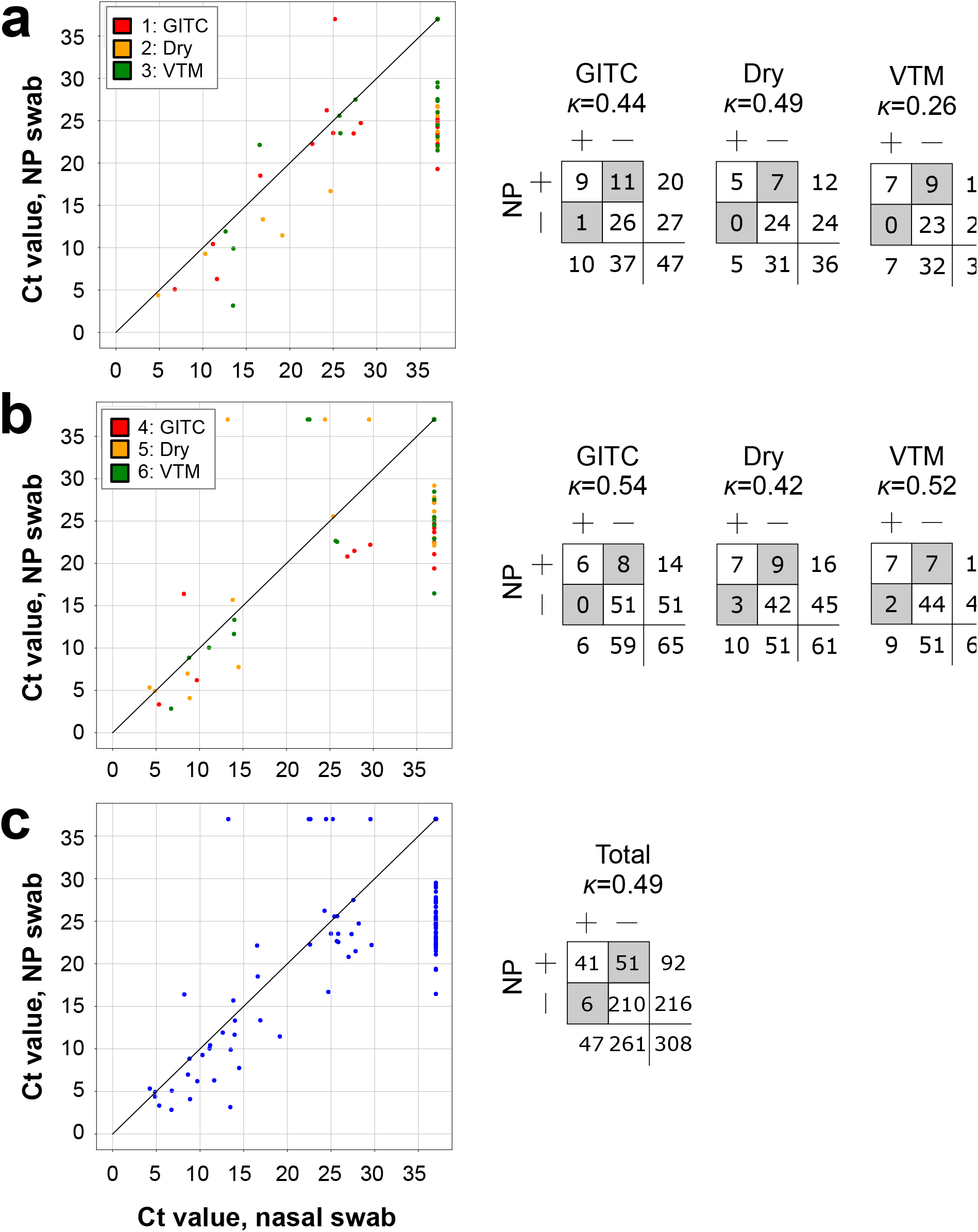
Comparisons of nasal-swab specimens to NP-swab controls. (a) Procedure 1; (b) Procedure 2; (c) total. Negatives are plotted with a Ct value of 37 (maximum cycle no.): vertically stacked data points at the far right of the plots (x-axis valu 37) are false negatives, while data points at the top of the plots (y-axis value = 37) are fal positives. Legend: study-arm numbers are the same as in Tables 1 and 2. *K*, Cohen’s ka

Our overall finding of low concordance was in contrast to some previous reports, which have found nasal-swab collection to exhibit excellent sensitivity as well as Ct-value concordance^13,15^, but was consistent with others^10,14^, including, for example, one recent study at a New York, USA, hospital that also noted lower nasal-swab concordance for higher Ct values^16^. Close review of these previous reports revealed that they differed in the type of specimen and/or result they used as a reference (e.g. any test-sample positive versus using NP swabs as the gold standard) and in the parameters they used in order to describe test performance (e.g. positive percent agreement versus sensitivity). To control for at least the latter, we extracted 2×2 contingency-table data from these reports to facilitate comparison to each other and to our own results (Table 3). Notably, many of these studies used a modified version of the CDC assay that did not report a LoD. Furthermore, of the studies that report the Ct values of their results, no viral-load conversion was provided, which is important since different RT-PCR assays and platforms have unique conversions between Ct value and viral load. Therefore, we were unable to systematically compare nasal-swab performance at low viral loads in these reports. These differences left open the possibility that inconsistent comparative performance of nasal-swab sampling might be explained largely by differences assay LoD, and possibly also by patient viral load. (We note that while the nasal-swab sampling protocols and transport media conditions varied between studies, our results suggest these differences are unlikely to affect detection).

**Table 3:** Comparison of nasal swab studies to date including their collection protocol and RT-PCR assay

| Study | Samples | Collection Method | Nasal +<br>NP + | Nasal +<br>NP - | Nasal -<br>NP + | Nasal -<br>NP - | Kappa | RT-PCR method (LOD in copies/mL) |
| --- | --- | --- | --- | --- | --- | --- | --- | --- |
| Berenger et al. (preprint) | 36 previously positive patients tested an average of 4d prior | APTIMA Unisex Collection kit (Hologic Inc.) used to swab both nares to a depth of at least 3 cm (or until resistance felt) and rotated 3 times | 22 | 2 | 5 | 7 | 0.53 | Alberta Public Health Laboratory (Provlab) (LOD: |
| Minich et al. (preprint) | 10 patients admitted with COVID-19 verified by NP swab | Sterile polyester head, plastic shaft dry swab inserted into one nostril to a depth of ~2-3cm and rotated for 5-10 seconds. Then placed in collection tube containing 0.5 – 1mL 95% ethanol and stored on dry ice | 3 | 0* | 0** | 1 | 1.00† | CDC protocol adaptation: 4µl RNA template, 100r 200nm probe, 3µl TaqPath, and water to a 10µl r Bio257 rad CFX384 Touch Real-Time PCR Detectic |
|  |  | Non-sterile cotton head, plastic shaft dry swab, same protocol as above | 3 | 0* | 1 | 2 | 0.66 |  |
|  |  | Non-sterile cotton head, plastic shaft dry swab, consumer grade | 4 | 1 | 1** | 1 | 0.30 |  |
| Wehrhahn et al. | 236 Australian patients tested at outpatient locations | Nasal swabs were inserted as far as comfortably possible and at least 2-3 cm inside one nostril, rotating the swab 5 times and leaving in place for 5-10 seconds. | 17 | 0 | 0 | 219 | 1.00† | Allplex™ 2019-nCoV Assay (Seegene, Seoul), Sout RT-PCR Detection Systems (no LOD listed) |
| Kojima et al. (preprint) | 45 adults that were recently tested for SARS-CoV-2 via standard NP swab testing | Supervised self-collected nasal swab used a CLASSIQSwab™ that the patient was instructed to insert into one nostril to the depth of 3-4cm and rotate for 5-10 seconds before storing the swab in RNA storage media (DNA/RNA Shield, Zymo Research Corp) | 19 | 4 | 4*** | 16 | 0.63 | Modified CDC assay with addition of N3 target to run on CFX 96™ Touch or Connect Detection Syst |
| Tu et al. (preprint) | 498 individuals tested at 5 different ambulatory centers | Nasal swabs collected with a foam swab (Puritan 25-1506 1PF100) via inserting in the vertical position into one nasal passage until gentle resistance and leaving the swab in place for 10-15 seconds and rotating. Swabs were stored in viral transport media. | 47 | 1 | 3 | 447 | 0.96 | Quest Diagnostics SARS-CoV-2 RNA, Qaulitative R Capistrano, CA) targeting N1 and N3 (nucleocapsi |
|  |  | Mid-turbinate swabs collected with a nylon flocced swab (MDL NasoSwab A362CS02) via inserting in the horizontal position into the nasal passage until gentle resistance is met, leaving the swab in for 10-15 seconds and rotating | 50 | 2 | 0 | 452 | 0.98 |  |
| Basu et al. (preprint) | 31 nasal swabs tested out of 101 samples collected in an adult ED | Dry nasal samples were obtained with swabs supplied with the Abbott assay (Puritan Medical Products 25-1506 1PF100). Nasal samples were obtained from both nares. | 17 | 1 | 14 | 69 | 0.61 | Abbott ID NOW (LOD 125 genome equivalents/m Cepheid Xpert Xpress SARS-CoV-2 test (LOD: 250 |

\*1 nasal swab was inconclusive, while NP swab was negative, \*\*1 nasal swab was inconclusive, while NP swab positive, \*\*\*2
samples were negative due to quantity insufficient. †Undependable given the zeros.

We therefore revisited the trend we observed of a rise in nasal-swab-negative discordant results (false negatives) with higher Ct value. Recently we demonstrated that Ct values for the SARS-CoV-2 RT-PCR assay and platform used in the present study are reliable quantitative measures of viral load, and introduced a conversion from Ct value to viral load^6^. Building on those findings, here we asked what the concordance would have been, for our nasal-vs.-NP data, had the LoD of our assay been higher than its actual 100 copies/mL. Specifically, we re-calculated kappa for different LoD cutoffs, and found that kappa rose steeply from ∼0.5 (low concordance) to 0.8-0.9 (excellent concordance) as the LoD cutoff was increased from 100 copies/mL through 1,000 copies/mL and beyond (Fig. 3). This finding strongly supports the view that nasal swabs miss many if not most patients with low viral load (below ∼1,000 copies/mL), but is reliable for patients with medium or high viral loads, potentially resolving disagreements among previous reports.

**Figure 3:**
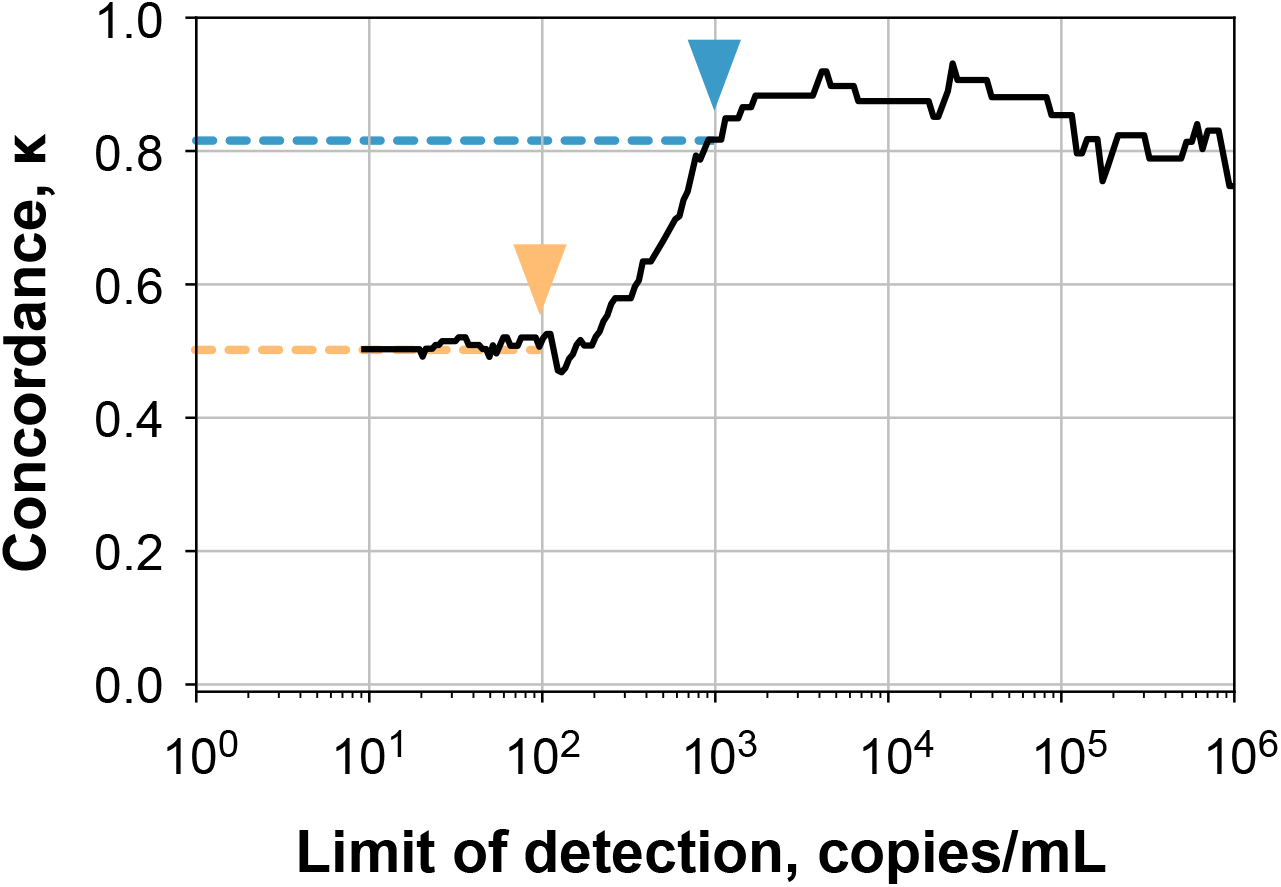
Low vs. high concordance depends on low vs. high assay sensitivity. Concordance (measured by Cohen’s kappa) plotted against assay LoD. With LoD of 100 copies/mL (yellow arrowhead) the Abbott assay detects false negatives in nasal-swab samples, resulting in low concordance (0.49; yellow dotted line). An assay with LoD of 1,000 copies/mL (blue arrowhead) would have missed these false negatives, which would have yielded a high observed concordance (0.82; blue dotted line).

## Discussion

It is widely acknowledged that resolving the damage that the COVID-19 pandemic has wrought to health, the economy, and society will require distributing and scaling up testing to unprecedented levels. For this reason, there is great and widespread interest in developing alternatives to NP-swab sampling for COVID-19 diagnosis. Governments and medical institutions alike have expressed interest in adopting nasal swabs as an alternative, as the self-administration of these swabs would allow vastly increased testing capacity, save PPE, and ease the burden on healthcare workers. Independently, the ability to transport swabs to testing locations without need of transport media such as VTM would further streamline testing processes. Reflecting this interest in nasal swabs, the US CDC has removed the “preference” specification for NP swabs from their interim guidance and note that nasal swabs are an acceptable alternative specimen as of 29 April 2020^4^. However, confidence in population-scale testing strategies based on nasal swabs is complicated by conflicting reports as to how well they perform relative NP swabs, the antecedent gold standard, as we have described.

Our results strongly suggest that concordance between nasal and NP swabs depends on the LoD of the PCR assay used to measure positivity, with high concordance for medium-to-high viral loads and low concordance for low viral loads, which may lie below the LoDs of various assays currently in widespread use (Fig. 3)^6^. We find that nasal swab samples reliably detect patients with viral loads ≥1,000 copies/mL but miss many patients who have lower viral loads, the majority in our study^6^. A complete biological explanation will require further study; however, one possibility is that in cases of high viral load, replicating virus may be more likely to spread to respiratory epithelium bordering and/or in the deeper portions of the anterior nares, where it can be recovered by nasal swab.

our findings may reconcile disagreements in prior reports, which have compared nasal swab performance only as a function of Ct values which are not comparable from study to study, not viral load, as we have done here,. We hypothesize that the outpatient/urgent care testing sites in these studies may have selected for patients early in the course of disease, when viral load is high^13,15^. For example, one study^13^ that showed high concordance used an assay with a negative Ct cutoff of 40, and only a handful of patient samples had Ct values above 35.

Although again the viral loads were not reported, the discrepancy between cutoff and Ct values suggested preferential sampling of patients with only high viral loads. (Note that a Ct value of 35 can correspond to different viral loads in different assays and the LoD of this assay was 4,167 copies/mL, over 40 times the cutoff in the assay we used.) Notably, the patient population in the present study consisted of both first-time and individuals with repeat testing for test of cure.

Many of the latter have been observed to exhibit low-level viral load for multiple weeks in the absence of severe symptoms and enrichment of these patients may impact the overall performance of NP and nasal swabs in individual studies. In other studies, such differences may be more or less obscured depending on the limit of detection of the assay in use which may straddle the 1,000 copies/mL threshold we found for nasal swabs to consistently detect infection.

Interestingly, we found no difference among transport media conditions and between sampling protocols, suggesting that lower sensitivity of nasal swab sampling is an overall limitation of the anatomical location of nasal swabs and that the protocols and media conditions we tested are interchangeable. Thus, for patients above a critical threshold of 1,000 copies/mL (Fig. 2), nasal swabs collected in VTM, GITC transport medium, and as dry swabs are all likely to perform equally well in the population, providing multiple potential options for specimen acquisition.

Our results suggest several settings in which nasal swabs may and may not best be used. Peak infectiousness is likely to occur near or shortly before symptom onset^20,21^ and nasopharyngeal viral load is often undetectable a week after symptom onset^2^. Lower-sensitivity testing would therefore likely miss patients with early developing presymptomatic infections and patients presenting multiple days after symptom onset. Notably, for those presenting later to care, a false-negative diagnosis could bear significant clinical implications in not only erroneously reassuring the patient and clinical team, but also excluding them from potentially useful and rationed therapies such as remdesivir^22^ or others. Importantly, based on viral load distribution in first-time tested individuals at our institution, ∼20% of newly presenting SARS-CoV-2 positive individuals would be missed if sampled solely using nasal swabs^6^, highlighting the potential magnitude of this problem.

Nevertheless, nasal swabs provide considerable advantage in terms of ease of collection and potential self-collection. Based on our results, they would serve best in high-test-volume, point prevalence screens in healthy populations, for example, in businesses and universities, where identification of highly infectious individuals will be a prelude to targeted testing with the most sensitive techniques possible to quell outbreaks and forestall local spread. Conversely, nasal swabs should not be used for screening symptomatic and especially hospitalized patients, where the more sensitive and resource intensive nasopharyngeal sampling would be justified, and help direct care and most appropriate use of infection control resources. In summary, whilst nasal swabs are a welcome addition to the armamentarium of tools needed to combat COVID-19, we should be well aware of possible limitations in diagnostic sensitivity and use this resource judiciously.

## Data Availability

Data, scrubbed of protected health information, will be made available upon request.

## Acknowledgements

The authors would like to acknowledge Dr. Yuan-Po Tu for graciously sharing his nasal swab protocol and Susan E. Devito, Kelly Gibbs, Angela E. Martignetti, Nancy R. Sheridan, Carl A. Santiago, Erin Martin, Erin K. Martin, Janet Mulvey, Mallory Flynn, Shannon Healy, Lauren Kennedy, Jill Lococo, Jose A. Abrego, Tracy Flynn, Jean M. Pietrantonio, Kelina Orlando, Carol Daugherty, (Carl) Adrian Santiago, Ann Marie Zampitella, Scott Hamelin, Maria Ortega, Frantzia Clermont, April Gagnon, Elvia Pires Barbosa Fernandes, Brenda Pinto, Brittany Williamson, Mairie Taline Dorcely, Sheila Moriarty, Kathy Corley, and Paula Stering for their assistance in conducting this study.

Abbott multi-Collect Specimen Collection Kits were a generous gift of Abbott Laboratories.

K.E.Z. was supported by a National Institute of Allergy and Infectious Diseases training grant (T32AI007061).

This work was conducted with support from The Harvard Clinical and Translational Science Center (National Center for Advancing Translational Sciences, National Institutes of Health Award UL 1TR002541) and financial contributions from Harvard University and its affiliated academic healthcare centers. The content is solely the responsibility of the authors and does not necessarily represent the official views of Harvard Catalyst, Harvard University and its affiliated academic healthcare centers, or the National Institutes of Health.

